# “You have to add your own character”: Medical Student Perspectives on Observing Clinical Encounters

**DOI:** 10.1101/2023.01.11.23284447

**Authors:** Rachel Stork Poeppelman, H Barrett Fromme, Rachel Yudkowsky, Matthew Lineberry, Laura Hirshfield

## Abstract

**Introduction:** The authors aimed to investigate how medical students interpret the observation of a more experienced clinician modeling both exemplary *and* flawed behaviors as well as how that interpretation influences their subsequent clinical performance.

**Methods:** We presented a recorded patient sexual history to 11 medical students. The recording displayed both exemplary and flawed behaviors. Students then obtained a sexual history from a standardized patient themselves. Using an interview methodology and constructivist analytic approach, we explored the process of learning from clinical observations.

**Results:** Students attended to flawed modeled behaviors, challenges specific to the task, and how areas of their own personal development were accomplished. They took a piecemeal approach to classifying modeled behaviors as done well or poorly based on previous instruction, experience, or perceived downstream effects. When applying their observations, students choose to copy, adapt, or avoid modeled behaviors based on their classification of the behavior.

**Discussion:** To optimize learning from observation, faculty can identify task-specific challenges and a student’s personal goals, which naturally draw the student’s attention, before observation in order to develop a shared mental model. When debriefing observed encounters, faculty may consider natural targets of learner attention, challenges specific to learning from observation, and factors likely to influence a learner’s judgement of modeled behaviors.

## Introduction

Medical trainees begin learning in the workplace by observing more experienced clinicians modeling their future role, a necessity for practical and patient safety reasons(1). However, descriptions of role modeling suffer from the lack of a unified definition. Existing definitions can generally be divided into two groups: those which focus on role models as admired, exemplary individuals and those which focus on the process of modeling knowledge, skills and attitudes for learners(2–5). Here we focus on the latter; in other words, role modeling as the process of teaching and learning by example(2,6–8).

Much of the research on the process of role modeling builds on Bandura’s social learning theory, which consists of four stages. The learner 1) observes what is modeled, 2) creates a mental representation of it, 3) practices what was modeled, and 4) is motivated to continue this practice through reinforcement(2,9,10). Learners make conscious decisions to imitate some behaviors and avoid others, taking pieces from different observed encounters(2–4,6,7,11–15). They make these decisions based, in part, on the perceived consequences of the modeled behaviors(2,11). Experts have called for role models to influence these decisions by debriefing with learners. Role models should reflect on the observed encounter in order to “make the implicit, explicit” and optimize learning(2,3,6,11,13,16–20). Learners also identify new goals and learning needs for themselves by recognizing things the model does particularly well(4,21). In contrast to this conscious process of adopting behaviors or goals, trainees also adopt observed behaviors and attitudes in an unconscious manner, a process which has proved challenging to study(3,12,16,22).

The existing literature analyzes a collection of experiences with multiple varied observations over time. No previous studies examine how trainees learn from observing a single, controlled clinical encounter. Additionally, previous research on the process of role modeling focuses primarily on positive role modeling, yet the educational value of observing both exemplary and flawed performance has been well described for motor tasks(18,23) and has also been reported for communication skills(17). Students recognize that exposure to mixed performance is important(24) and most day-to-day clinical encounters model both exemplary behaviors and behaviors which could be improved(7,16,25–33). Thus, examining how trainees learn from observing a mixture of exemplary and flawed behaviors is important to better approximate the real world.

We build upon existing research through an inductive exploration of trainee cognition and practice after observing a mixture of exemplary and flawed behaviors modeled in a single, controlled encounter. By focusing on a single, controlled encounter, we shift our focus to a deeper examination of the learning *process* rather than the range of knowledge, skills, and attitudes modeled in the workplace. In turn, a better understanding of how trainees process an observed mixture of behaviors in a single encounter and how they apply this in practice can help medical educators to optimize learning from observation. Such an understanding could influence the structure or debriefing of observed encounters.

## Methods

To investigate the effect of observing a mix of exemplary and flawed modeled behaviors, we conducted a video-based intervention with 11 medical students. Students observed a video-recorded mixed example history before taking a sexual history from a standardized patient themselves. Using semi-structured interviews, we explored trainee cognition proximal to observation of and practice with a standardized clinical encounter. This method contrasts with most previous research, which has relied primarily on trainee or educator recall of remote, diverse experiences with learning from observation. Here, we aimed to capture the range of student reaction to a common experience, to add new insights about learning from observation while at the same time, limiting recall bias and controlling for differences in observed encounters.

### Participants and Study Setting

Pre-clinical medical student volunteers were recruited from two medical schools (University of Chicago and University of Illinois at Chicago) over email by the PI (RSP). Purposive sampling from a large public allopathic medical school and small private allopathic medical school was selected to broaden the diversity of training experiences represented in the sample. Participation was incentivized with a $10 gift card to Starbucks. The study took place at the standardized patient simulation center at each institution and was deemed exempt by the IRB at each institution.

### Data Collection

Students watched a video of an attending physician (“Dr. Smith”) exhibiting a mixture of exemplary and flawed behaviors while taking a sexual history from a male patient (Appendix A). The behaviors modeled were selected, reviewed and revised by the authors, based on literature on best practices in sexual history-taking. Sexual history was selected because many students find it challenging. The attending in the video is a female faculty member at University of Chicago.

Participants were then recorded taking a sexual history from a female standardized patient (Appendix B)(34). The standardized patients debriefed participants on their performance. After the SP encounter, participants completed a survey. In the survey, participants evaluated the attending’s performance in the video using a global rating scale and checklist of communication behaviors and sexual history tasks, adapted from the Kalamazoo Essential Elements Communication Checklist and clinical guidelines^36-42^ (Appendix C). General demographic information was also collected from participants.

The PI (RSP) then interviewed the students about their experiences guided by an interview protocol (Appendix D) and the completed participant survey. The timeline of participation is depicted in Figure 1.

**Figure 1.**
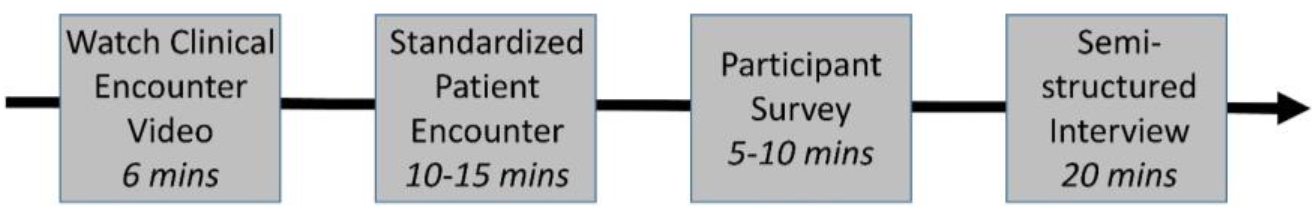
Participant Timeline

### Data Analysis

We conducted our analysis using a constructivist framework, which posits that there is not one fixed reality but multiple realities constructed from each individual’s interpretation of the world, in order to capture the range of student experience(35). In line with this approach and our research goal of conducting an exploratory analysis, one coder (RSP) analyzed the data, with input from the senior author (LH). We used an inductive analytic approach consisting of open-and-focused coding(36). Although existing theories were not used to create codes, the data is presented in the context of Bandura’s social learning theory to better situate the findings in the existing literature. The PI (RSP), a female graduate-level medical trainee at the time of analysis, performed the primary data analysis. The PI began by immersing herself in the data with a comparison of interview recordings to the professional transcription, to ensure accuracy. She coded the first five interviews with an open-coding approach to develop a codebook and summarized findings in an initial memo. She then used a narrower, focused set of codes to revise her analysis of the first five interviews and analyze the remaining six interviews. She composed an extensive integrative memo, inclusive of all data, to organize the codes around themes and patterns(36). Themes and codes from the interview analysis were supplemented by an interpretive content analysis of the video-taped SP encounters, which assumes that “meaning is not simply ‘contained’ in the text” and places a greater emphasis on interpreting the data in context.^34,35^. By comparing the SP encounters to the observed clinical example, we were able to assess for similarities in phrasing or movement suggesting the influence of Dr. Smith’s performance on the student’s performance.

All memos were shared and discussed with the senior author (LH) throughout this process to ensure analytic rigor. The richness of the 198 minutes of participant interviews, supplemented by the recorded standardized patient encounters, provided sufficient information power to complete this exploratory analysis of medical student cognition.(37)

## Results

### Demographics

During the spring of 2017, eleven medical students were interviewed. Five were first-year medical students and six were second-year medical students. All students had previous lecture-based instruction on taking a sexual history and had experience taking a sexual history from a standardized patient. Two students had additional experience in a free clinic. Of note, although we intended to recruit both male and female participants, our sample consisted only of women. Women tend to volunteer more for surveys/studies, which may have impacted the gender composition of our sample(38).

### Findings

Our findings are organized in three stages of the process of learning from observation: 1) learner attention, 2) judgement of modeled behavior, and 3) learner application. This 3-stage process, which echoes the first 3 stages of Bandura’s social learning theory, provides a framework through which we can better understand and describe the process of learning from observation. The fourth stage of Bandura’s theory, motivation, is not addressed as the reinforcement of modeled behaviors is beyond the scope of this work.

#### 1. Learner Attention

The focus of learner attention in an observed encounter has been identified as an important determinant of learning(39). Students described four determinants of learner attention in observed encounters: 1) flawed modeled behaviors, 2) behaviors addressing task-specific challenges, 3) behaviors relevant to personal goals, and 4) task-specific history content. Students describe poorly performed behaviors as sticking in their minds, sometimes to the point of distraction.

> *“[I remember] Flinching really hard when she asked, ‘How many women do you have sex with?’ I was like ‘No! you can’t do that!’ That stood out to me pretty aggressively*.*”*

Students also focused their attention on tasks they anticipated to be particularly challenging. For example, in taking a sexual history:

> *“I always want to pay attention to how the doctor asks-you know asking about sexual partner number is difficult*… *I’m always looking for different examples of how to ask that. [I pay attention to] how the question is phrased because I think that’s hardest for me, how to phrase it. “*

If a student had a specific personal goal for their standardized patient encounter, their attention was drawn to how the clinician accomplished that goal, either as a template or counterpoint. Students also uniformly reported paying attention to the content of the sexual history in the video, as a review of the kinds of questions they should ask in their history. Notably, a few students reported that their attention to the thoroughness of the history made them nervous about missing pieces of information in their own SP encounter. This contrasts with findings from studies of peer observation, which indicate students derive self-efficacy from observing peers perform well(40).

> *“ Usually, I am pretty confident taking a sexual history*…*But I think [the video] made me second guess some of the information I needed to take down*.*”*

Overall, students’ attention was drawn to four things in the observed encounter. Often, students saw these attention grabbers as useful, but in some cases, they were distracting or decreased a student’s self-efficacy.

#### 2. Judgement of Observed Behavior

The second stage in learning from observation is the creation of a mental representation of what was modeled, requiring the learner to categorize what was observed and make connections with previous experience. Our findings echo previous descriptions of a piecemeal approach to interpretation of the observed clinician’s performance(6,12–14). Students valued the individual behaviors independently, rather than applying an overall judgement to the entire encounter.

In our study, a student’s approach to determining the merit of an observed behavior varied between a pattern-based categorization and a logic-based approach. When observed behaviors clearly paralleled or contrasted with a student’s existing habits, they were automatically classified based on those habits. For example, several students automatically classified Dr. Smith’s assumption of the patient’s partner’s gender as a flawed behavior due to their own practice of asking about gender before using a gendered term.

> *“Something I noticed from the video that kind of bothered me was the fact that the doctor kind of assumed too much about whether the patient was with a male or female or both. She immediately assumed that he had a female partner. When I ask the questions, I ask, “Have you been with men, with women, with both?” to make sure you’re inclusive in that sense, and that has really been emphasized in our class, and I think that’s really important*.*”*

Alternatively, in the logic-based approach to classification, students based their reasoning on previous instruction, experience, or a consideration of downstream effects.

The estimation of downstream effects was often anchored to students’ observations of the patient’s reaction to the behavior.

> *“I was honestly surprised that the patient wasn’t more taken aback when he was answering that question [about HIV testing]. He seemed pretty much to roll with it, and I was like, ‘Well, I guess it’s fine*.*’”*

Students were sometimes uncertain how to classify behaviors due to a knowledge deficit, lack of student confidence, or observation of an inherently ambiguous behavior.

> *“[When I said she could have improved her] slang, I guess that one argument is that you shouldn’t say ‘Getting it up’, which is why I put that. That’s not my personal feeling, so I don’t know, maybe I differ from the correct way. Maybe I should be corrected. I feel like the patient says something and that’s what they’re comfortable with, to me, it’s not a bad thing*.*”*

Interestingly, the classification of behaviors was not influenced by a pre-existing relationship with the clinician in the video. All students described her performance as mediocre or good with room for improvement. However, several students acknowledged recognizing that the clinician was “acting”, which may have negated the influence of a pre-existing relationship. Additionally, when students failed to categorize a flawed behavior as such, they often were cognizant that the behavior was performed differently than how they would approach it but attributed this difference to either personality differences in communication or to the absence of the time pressures of the attending role.

> *“I feel like the style she had is … like time-crunch medicine, like real, real medicine. Like you’re in the office, you got to get things done* … *right now, I don’t relate to that because I’m a student, and I have all the time in the world*.*”*

#### 3. Learner Application

After the learner creates a mental representation of their observation, they practice what was modeled(10). In our study, students recognized four ways that watching the video influenced their practice with the standardized patient: 1) copying modeled behaviors, 2) adapting modeled behaviors, 3) avoiding modeled behaviors, and 4) as a reminder of salient history content. When copying modeled behaviors, students reported trying to copy both specific phrasing and more general communication behaviors.

> *“Her being able to say ‘I’m really sorry’, ‘that seems really hard’, ‘that seems really hard for you’… I took note of that too, and wanted to – I remember mentally noting like if something happens, that’s good language to use*.*”*

Previous research describes students emulating models they see as similar to themselves(12,14). Most students saw Dr. Smith as dissimilar to themselves, both because of the flawed behaviors modeled and differences in age, experience, and demeanor. Despite this, all students described copying some behaviors from the video. Previous work describes adaptation of behaviors as subsequent to copying(12). However, in our study students described adapting observed behaviors that either didn’t fit well with their personal communication style or that they thought could be improved upon immediately following observation, instead of copying first.

> *“[I wanted to replicate] the tone of the questions, her overall empathy and building rapport with the patient. Beyond that, I mean you have to add your own character to your interactions. I’m not going to copy her. She’s herself*.*”*

Students found that flawed modeled behaviors reinforced things to avoid in their own encounters.

> *“I think reacting the way that I did to some of the things that weren’t done well made me very aware to not do them in my own encounter*.*”*

In terms of practice challenges, some students reported struggling to either copy or adapt the behaviors they observed in a new context.

> *“Initially, I was relying on the video as a template and then once I realized I was going to be interviewing a woman I was like, ‘Okay, well, this is a little different*.*’ “*

Additionally, relying on the video as a template resulted in incomplete histories from a few students who failed to recognize that portions of the sexual history were missing from the video.

Overall, students described that the observed encounter was a smaller influence on their performance with the standardized patient than previous experience and instruction. However, in addition to the behaviors students consciously copied, adapted, or avoided, students also acknowledged that the video likely had an additional subconscious influence on their performance.

> *“I think even if it’s subconscious, there has to be a little bit of mirroring. Like you just see it. It’s one of our preceptors. It’s someone that we – or I – feel like I’m supposed to be learning from. I think even consciously and subconsciously I take things that she does*.*”*

## Discussion

We set out to further explore the process of learning from observing the mixed performance of an experienced clinician in a single encounter. The process described by students in this study echoes and expands on three of the subprocesses outlined by Bandura’s theory of social learning: attention, retention, and reproduction(10). Our results have implications both for the creation of instructional videos and for trainee learning from clinical observations, as summarized in Table 1.

**Table 1:** Implications for Structuring Observations

| Key Take Home Points | Implications- Instructional Videos | Implications- Clinical Observation |
| --- | --- | --- |
| While observing clinical encounters, learners pay attention to:<br>- Flawed modeled behaviors<br>- Behaviors relevant to learner-specific goals<br>- Behaviors addressing task-specific challenges<br>- Task-specific history content | *\*Before creating videos*, identify key take home points to be embedded as flawed behaviors, learner goals, or task-specific challenges (draw learner attention) | *\*Before observed encounters*, identify learner goals and specific challenges anticipated in order to direct trainee & model attention<br>*\*After observed encounters*, focus debrief on behaviors trainees attended to or behaviors relevant to learner goals/task-specific challenges |
| Learners classify observed behaviors in a piecemeal fashion based on patterns or logic influenced by:<br>- Existing habits<br>- Previous instruction<br>- Theorized downstream effects<br>- Observed patient reaction | *\*Before creating videos*, investigate prior instruction on the topic to anticipate a student’s interpretation of instructional videos<br>*\*When creating videos*, use the patient’s reaction to a behavior displayed to reinforce behavior classification as positive or flawed | *\*After observed encounters*, shape the debrief by:<br>1) Probing trainees for relevant previous instruction/habits<br>2) Highlighting unusual patient reactions to limit their influence<br>3) Framing behaviors in intended downstream effects |
| Learners may be uncertain how to classify a behavior due to lack of knowledge or confidence | *\*After observed video encounters*<br>1) Gather reactions: did anything surprise or puzzle the learner?<br>2) Ask the learner how any learner goals or specific | *\*After observed encounters*<br>1) Gather reactions: did anything surprise or puzzle the learner?<br>2) Ask the learner how any learner goals or specific challenges were accomplished and probe for knowledge deficits |
|  | challenges were accomplished and probe for knowledge deficits<br>3) Leave time for questions | 3) Leave time for questions |
| Challenges for some learners<br>- Transfer (Context specificity)<br>- Recognizing omissions<br>- Negative self-efficacy | *\*When creating videos,* considering displaying peer performance (rather than supervisor) to improve trainee self-efficacy | *\*After observed encounters,* highlight generalizable behaviors and omitted behaviors<br>If appropriate, acknowledge that observing can sometimes be intimidating for learners. |
| Flawed behaviors can distract learners from attending to events for a period of time after observing the behavior | *\*When creating videos,* do not portray key take home points immediately following flawed behaviors |  |

Raîche et al. described the importance of directing learners’ attention in observed encounters(39) but not how attention is naturally directed. Horsburgh et al. found that learners paid attention to behaviors that align with traits they value in a doctor(2). In contrast, we found learners attended to flawed behaviors, behaviors addressing task-specific challenges, behaviors relevant to learner-specific goals, and the overall history content. The tendency of flawed behaviors to attract learners’ attention may reflect the negativity bias, our innate tendency to both attend to and learn from negative stimuli more readily than positive stimuli (41).

Previous works show the interpretation of observed behaviors to be influenced by the observed response of the patient(2) and have related that to Bandura’s vicarious reinforcement(10), but not the learner’s approach to this interpretation. Students in our study described two approaches to deciding the value of their observations: a pattern-based classification and a logic-based classification. We also found that there were observed behaviors some trainees had trouble classifying, either due to lack of knowledge or confidence. We know that reflection, a process which enables learning from experience, varies among trainees and is a skill which can be developed(19). The factors which influence a student’s ability to learn from observing another’s experience likely parallel these, however that is beyond the scope of this work.

Despite the previous finding that students emulate models they see as similar to themselves(12,14), we found students still copied behaviors from a model they perceived as dissimilar due to age, experience, and demeanor. We also found that learners adapted behaviors to improve upon them or to better suit their personal communication style. This differs from previous works which either do not describe adaptation or describe it as occurring after copying(12). This may reflect a difference between learning motor skills and communication skills, which are inherently more personal skills, and thus unlikely to be copied exactly.

Experts have advocated “making the unconscious conscious” for trainees through debriefing clinical encounters(16). Our findings can help guide clinicians to optimize observed encounters and instructional videos by shaping instructional videos, debriefing of both videos and encounters, and highlighting the importance of setting up a shared mental model before observation, as summarized in Table 1. Many debriefing frameworks begin with a “reactions” phase(42,43). Similarly, we suggest role models or educators displaying an instructional video begin a debrief with asking the learner if anything they observed surprised or puzzled them. In addition to those “reactions”, the behaviors likely to draw the learners attention (described above), specific challenges to learning from observation, and factors likely to influence behavior classification, are good topics for debriefing. Role models may also want to acknowledge that some learners will feel intimidated by observation and mitigate this effect with reassurance about the learner’s trajectory. Educators may want to use peers in instructional videos to mitigate this effect. The importance of involving the learner in a discussion, rather than simply articulating teaching points to them should be emphasized(42). For example, without probing trainees for their interpretation of what was observed or leaving space for questions, learners may have lingering uncertainty or persistent knowledge deficits which could have been addressed in the debrief.

Limitations of this study include a small sample size consisting only of female students, which limits the ability to extrapolate our findings to male students. Additionally, students acknowledged a subconscious influence of observation on their own behavior, which was unable to be further characterized by our methods. Lastly, given previous findings that students emulate models they see as similar to themselves(12,14), our findings may have differed with a model the students saw as more similar to themselves. However, many students specifically identified the flawed behaviors displayed as a reason for their perception of dissimilarity, suggesting that observation of a mix of flawed and exemplary behaviors often results in perceived dissimilarity. This is a potential area of future investigation.

Overall, our findings about the process of learning from observation can help guide clinicians in making observed clinical encounters and educational videos more productive, specifically in designing instructional videos, helping trainees focus their attention and structuring a productive debrief.

## Data Availability

All data produced in the present study are available upon reasonable request to the authors

## Acknowledgements

The authors wish to thank the participants who shared their insights with us. We would also like to thank Kris Slawinski and Jenna Jackson at the University of Chicago Clinical Performance Center and Laura McKenzie at the University of Illinois at Chicago Simulation and Integrative Learning Institute for invaluable logistical support.

## Declaration of Interest Statement

The authors report there are no competing interests to declare.

## List of Appendices

Appendix A. Video Script

Appendix B. Standardized Patient Encounter

Appendix C. Participant Survey

Appendix D. Interview Guide

## Notes

### Competing Interest Statement

The authors have declared no competing interest.

### Funding Statement

Funding for participant incentives, standardized patients and transcription services was provided by a pilot grant from the Bucksbaum Foundation for Clinical Excellence at University of Chicago.

### Author Declarations

Approved by the IRB at University of Chicago and University of Illinois at Chicago

