## Appendix A for "“You have to add your own character”: Medical Student Perspectives on Observing Clinical Encounters"

### Appendix A. Video Script

***Background information for patient role: Edward Stone***

*You are a 35 year-old man who is establishing care with a new primary care physician. You have been having trouble with erectile dysfunction and difficulties climaxing during intercourse. You are stressed at work and 8 months ago you adopted a 5 m/o baby girl, who is now 13 months old.*

*Appearance- You are a healthy male. You are well groomed, wearing office casual clothes. You have a closed posture for the majority of the interview. Your hands are lightly clasped in your lap most of the interview. You use your hands to talk a lot.*

*Affect/Behavior- You are obvious worried about your erectile dysfunction. You are embarrassed by your situation but cooperative and pleasant. There is a sense of sadness or depression in your demeanor. You are restless and withdrawn. When talking about how sexual problems are affecting your relationship, you will break normal eye contact and look down each time you talk about either the sexual dysfunction and subtly switch the position of your feet/legs- particularly when you feel uncomfortable. Your voice is soft but clear towards the beginning of the interview but will return to normal loudness as you become more comfortable.*

*The doctor is seated at a desk in front of a computer, the patient is seated on the exam table in a gown. Both parties are visible in the camera frame.*

**Doc**: As a part of your full physical and as a new patient, I would like to ask you some questions about your sexual history. I ask these of all of my patients and they are important for me to help you stay as healthy as possible. Anything we discuss will of course remain confidential between me and you. Is that ok?

**Patient**: Yes

**Doc:** Are you sexually active?

**Patient:** Yes

**Doc:** How many women have you been sexually active with in the last 3 months?

**Patient:** Just my wife

**Doc:** How long have you and your wife been together?

**Patient:** 10 years this summer.

**Doc:** Are you or your wife having any sexual difficulties?

**Patient:** Well… this is embarrassing *(Shifts his weight, looks down at his hands)* … lately I haven't been able to … you know… when we're together.

**Doc:** Could you be more specific?

**Patient:** You know… get it up. And when I do, I … I can't always finish.

**Doc:** Oh, well is it more of an issue getting it up or keeping it up.

**Patient:** More-so trouble with keeping it up. I'm a little embarrassed to be talking about this with someone I'm just meeting today. I know that people say it is common but you never think…

**Doc:** *(interrupts)* When did you first notice this?

**Patient:** Hmm. Probably around the time I started my new job at Martin & Martin. I was promoted to project manager about 6 months ago- it's a great opportunity for my career but it came with a much bigger workload. I was pretty stressed out with the transition and with our new baby. She really isn't…

**Doc:** *(interrupts)* New baby? Congratulations! Boy or girl?

**Patient:** Girl. Her name is Lillian, we adopted her about 8 months ago. We're so happy to have been able to adopt her, but it was a lot of change and added responsibility all at once. Whenever I'm stressed out with work or with the baby, it does seem to make things worse in the bedroom. When all of this started, I blamed it on the Prozac I was taking at the time but I have been off of that for months and the problem has only gotten worse, not better.

**Doc:** Is there anything that helps?

**Patient:** It seems like if I've had a few drinks beforehand that I don't have as much trouble getting it up. I think it calms my nerves and I am more able to focus on my wife.

**Doc:** Other than your job and the new baby, do you have any other new stressors in your life?

**Patient:** Work is the main thing stressing me out right now. The baby just adds a level of sleep deprivation to the challenges at work. Though I guess all of this hasn't been great for my marriage.

**Doc**: Has there been any change in your sexual desire with these issues?

**Patient:** I wouldn't say there has been a change in desire, but I do get nervous about performing. Again, that's why I think the alcohol helps.

**Doc:** How many times a week do you drink alcohol?

**Patient:** Maybe once or twice. Like I said, my job and the baby keep me pretty busy.

**Doc:** On average, how many drinks do you have when you drink?

**Patient:** Maybe 2.

**Doc:** Do you use any drugs?

**Patient:** No of course not. I'm a father.

**Doc:** What kinds of sexual practices do you and your wife engage in? Vaginal? Oral? Anal?

**Patient**: Umm, primarily vaginal or oral. She's not really open to anal.

**Doc**: Do you use contraception or condoms?

**Patient**: No. We aren't able to conceive, we tried for years so there's really no point in contraception.

**Doc:** I'm sorry to hear that. That must have been hard for you.

**Patient:** It was. But now we have Lillian *(smiles warmly)* and I can't imagine things working out any other way.

**Doc**: I'm so happy for you. How many partners have you had in your lifetime?

**Patient**: *(looks up as if counting in his head)* Let's see… 5 total.

**Doc:** Did you ever have trouble with erections with any of those partners?

**Patient:** No

**Doc**: Have you or your partner ever been treated for an STI before?

**Patient**: What is an STI?

**Doc**: Sexually transmitted infection, like HIV or herpes.

**Patient**: Oh no, nothing like that.

**Doc**: Have you and your partner ever been tested for HIV?

**Patient**: Back when we first started dating. We both got tested and it came back negative so that's when we stopped using condoms.

**Doc:** Do you have any other concerns that we haven't touched on?

**Patient:** My main concern is my trouble with erections. I'm worried that my wife is becoming increasingly unhappy with our sex life, which is putting a strain on our marriage. I'm afraid she will leave me if I can't fix this and the more I think about that, the worse the trouble seems to get.

**Doc:** I'm glad we were able to discuss this today. Now that I have a little more background, let's talk about what we can try to help improve your sexual functioning.

*Camera fades to black.*
