## Appendix B for "“You have to add your own character”: Medical Student Perspectives on Observing Clinical Encounters"

**Appendix B. Standardized Patient Encounter**

### Danni Allen

**Chief Concern: To establish care with new Dr. and get Pap Smear**

**Recruitment Profile: Female, Age Range- 20-30**

#### Case Summary

You are a 24 year-old woman who comes to the clinic today with no chief complaints. You want a refill of birth control and to establish care with a healthcare provider.

#### Case Setting

You are in the outpatient clinic. This is your first visit to this clinic.

#### Case Challenge

The challenge for the learner is to practice their sexual history taking skills. The learners will be challenged to engage you in a non-judgmental and open manner.

#### How to Appear During the Encounter

General appearance/ grooming: You are a healthy young female. Your hair is clean and combed but not styled. You have make-up that is well applied.

Dress: You should be dressed in clean casual clothes.

Description of Affect and Behavior

You are a happy, outgoing young woman. You are cooperative, pleasant, relaxed and self-confident. Your speech is clear and easy to understand. You will maintain normal eye contact, a little less during the sexual history portion of the interview. You will respond to directed questions with information but only information asked. You enjoy your job very much. You do not know what is included in an “annual check-up”.

****When asked about your sexual history**

You will avert your gaze when talking about your sexual history and will become visibly uncomfortable when specifics about the more personal information is asked. If the student does not assure you of confidentiality before asking about your sexual history you should prompt them with a statement like “You aren’t going to talk to anyone about this stuff right?”.

You will answer straightforwardly but will initially not offer any sexual history without being asked.

If asked how many partners you have had in your lifetime, you will have to think a little bit and then state “I think it has been somewhere between 15 – 20.”

If the learner implies that you have been promiscuous, you will get defensive. The number of partners is normal to you since your girlfriends have all had 2-3 boyfriends every year. You have been monogamous with each of your partners. You do not feel that you are engaging in high-risk sexual behavior. If the learner approaches you in an open and accepting manner, you will be receptive to any sort of counseling. If you perceive that the recommendations are condescending or judgmental, you will get defensive and disengage.

If the encounter is drawing to a close and the student has not asked about partners, practices, contraception, pregnancy, previous STIs, STI testing or STI protection you may offer a prompt.

Prompts to use at the end of the encounter:

*If the student has not asked about previous STIs, STI testing and/or STI protection:*
Admit to the student that you are worried about whether or not you need STI testing but that you feel guilty about your worry because you are in a monogamous relationship and you don’t want it to seem like you don’t trust your boyfriend

*If the student does not ask about sexual abuse:*

State to the learner when they ask if there is anything else: “I just don’t want you to think I’m uncomfortable talking about this stuff because I’m in a bad relationship. Its just hard to talk about. My relationship is great right now, unlike some other guys I’ve been with”

#### Present Life

Age: 24

Date of Birth: Use your own birth date, 19___

Level of Education: You have a BS degree in biology.

Occupation: You are an assistant at a realty Company

Marital Status: Single. You have a boyfriend.

Hobbies/Activities: Hanging out with friends, going out to eat, tennis, going to the gym

Life Stressors: Sometimes work can get a little stressful

#### Reason for Visit (Chief Complaint):

***"I need a new doctor and I think I need a Pap Smear."***

#### History of Present Illness

Information relating to you primary complaint: **No Complaint**

| | |
|---|---|
| **Context** | • You have not seen a primary care provider since you were a kid. You have mostly been going to different free or Planned Parenthood clinics for refills on your birth control and the occasional cold. You just got health insurance through your job and your parents told you that you should see a doctor for regular checkups. |

| | |
|---|---|
| **Concerns** | • You don't have any concerns but wonder what you are supposed to have done as part of a "checkup".<br>** Risk Factors:<br>• multiple sexual partners<br>• unprotected intercourse<br>• oral, penile-vaginal, digital-vaginal, and anal receptive intercourse |

#### Past Medical History

| | |
|---|---|
| **Pt's response to "how is your overall health?"** | Your overall health has generally been pretty good. |
| **Obstetrical History** | • You started your periods when you were 12 years old.<br>• You have never been pregnant. |
| **Medications** | Birth control Pill (generic version of Alesse)<br>You have been on the same oral contraceptive since 14 years old. |
| **Most recent visit to a health care provider** | *Primary Care MD*: Here to establish with you<br>*Most recently:*<br>• Your last pap test was 1 year ago at Planned Parenthood and it was normal. You have had a few now (about every 2-3 years)<br>• When you were 14 year old, you started on birth control pills to help regulate your menses, which were heavy. |

| | |
|---|---|
| | You have primarily gotten healthcare through Planned Parenthood and other free women's health clinics over the years. |

#### Family History

| | | |
|---|---|---|
| **Father** | Alive<br>Age - 50 | Hypertension but otherwise healthy |
| **Mother** | Alive<br>Age- 49 | healthy, no major medical problems |
| **Siblings**<br>Brother | Alive<br>Age- 21 | healthy, no major medical problems |

#### Sexual History

| | |
|---|---|
| **Sexual orientation** | Heterosexual |
| **Sexually active at present** | Yes |
| **Number of current sexual partners** | Your boyfriend is your only current partner. You have been dating for the past 3 months. You don't think that he is with anyone but you. You don't know his sexual history. Together you practice oral (which you term "giving head"), vaginal and anal receptive intercourse and use condoms intermittently. |
| **Past partners in the past year** | You have had 2 other partners in the past year (both male). They were both ex-boyfriends (serial monogamy). |

| | |
|---|---|
| **Number of previous sexual**<br><br>**partners** | ***If the learner specifically asks about gender with this question you should answer with "I'm with a man right now" But if they assume gender do not offer any hints.***<br><br>*Around 15-20 partners*<br><br>• 3 were women—these were each one time experiments in college. It was mostly kissing, a little fingering (digital-vaginal intercourse), and some head (oral sex). If asked you have ever used dental dams, you state you don't know what that is. **Note: these were separate encounters with 3 different women**<br><br>• 10-15 Male partners—All have been monogamous relationships. There were all boyfriends who you dated, which mostly lasted 3-6 months on average. You used condoms most of the time but not always because you were on birth control pills. You practiced oral, vaginal and anal receptive intercourse.<br><br>• If you are asked about the first time you had sex you will answer, 17 yo as that was the first time you had vaginal sex.<br>• If you are asked about specific practices or previous partners you will describe the history from 15 y/o on. |
| **Age began to be sexually active** | Age- 15- you had a boyfriend when you were 15 but you only "gave him head" so you don't consider that "real sex". |

| | |
|---|---|
| **Sexually transmitted infections** | Chlamydia- age 17. It was treated with several pills and you have not had any STI's since. You have never been tested for HIV (if asked, you don't think that you should be concerned since you've only dated one person at a time). |
| **Sexual abuse history** | If asked you will shyly reveal that when you were a freshman in college, you dated your biology teaching assistant (TA) for a month. You felt pressured to have sex with him even though you didn't want to. You are not sure whether that is considered rape/sexual abuse or not. You are embarrassed that it happened and felt it was your fault because you were flirting with him. You are not supposed to be dating your TA anyway.<br><br>Since you don't really see the encounter with your TA as sexual abuse:<br>If you are **only asked "have you ever been sexually abused?** You will hesitate and state **"not really"** or **"I don't think so".**<br>If you are asked a more inclusive formulation of the question, such as "**Have you ever felt pressured to have sex when you didn't want to?"** or adds a normalizing statement, etc. you will disclose the information about the TA |

#### Menstrual History:

| | |
|---|---|
| **Menarche** | At 13, started birth control at 14 to help with heavy menses. |
| **Cycle** | Regular, every 4 weeks, lasts 5 days |
| **Menstrual symptoms** | Some cramping, go through about 3-4 pads a day, not particularly heavy, much better since you have been on birth control |

#### Personal Habits:

| | | | |
|---|---|---|---|
| Tobacco | Current: | Cigarettes | *Quantity*:<br>1-2 cigarettes daily when you go out with friends (you have smoked like this for 2 years). You are a social smoker and have not tried quitting. |
| Alcohol | Current: | Beer, wine or hard liquor | *Quantity:*<br>3-5 drinks per week |
| Drugs | Past: | Marijuana | *Quantity:*<br>Occasionally in college |
| Diet | regular well balanced diet, three meals per day | | |
| Caffeine use | minimal (1-2 cups of coffee/tea or cola per day) | | |
| Exercise | daily exercise<br>*Type of exercise* – gym and tennis | | |
