## Appendix C for "“You have to add your own character”: Medical Student Perspectives on Observing Clinical Encounters"

**Appendix C. Participant Survey**

*Unique identifier: First five letters of the name of your high school+ last 3 digits of your phone number*

_______________________________________________

Rate how well the physician in the video performed on a scale of 1 to 5 in the following areas (1= poor, 3= fair, 5= excellent).

| | | | | | |
|---|---|---|---|---|---|
| communication skills | 1 | 2 | 3 | 4 | 5 |
| content of sexual history | 1 | 2 | 3 | 4 | 5 |
| professionalism | 1 | 2 | 3 | 4 | 5 |

Think back to the video you watched. Please rate the physician's performance in the following areas based on your memory of the video-tape.

| | Not Done | Needs Improvement | Does Well |
|---|---|---|---|
| **Build a Relationship** | | | |
| Shows interest in patient as a person | | | |
| Uses words that show care and concern throughout the interview | | | |
| Uses tone, pace, eye contact, and posture that show care and concern | | | |
| **Open the Discussion** | | | |
| Asks "Is there anything else?" to elicit full set of concerns | | | |
| **Gather Information** | | | |
| Clarifies details as necessary with more specific or "yes/no" questions | | | |
| Summarizes and gives patient opportunity to correct or add information | | | |
| Transitions effectively to additional questions | | | |
| **Understand the Patient's Perspective** | | | |
| Asks about life events, circumstances, other people that might affect health | | | |
| Elicits patient's beliefs, concerns, and expectations about illness and treatment | | | |
| Responds explicitly to patient statements about ideas, feelings, and values | | | |
| **Sexual History** | | | |
| Assures the patient of confidentiality | | | |
| Uses a non-judgmental tone (e.g. as if talking about the weather) | | | |

| |
|---|
| Uses inclusive language (e.g. does not specify gender of partner unless patient does first) |
| “Normalizing” questioning technique (e.g. “ many of my patients have a history of STI, have you ever been treated for an STI?”) |
| Uses anatomical/professional terminology, not slang |
| Asks about:<br>STI risk factors (Partners, practices, protection) |
| Pregnancy (“trying”, contraception) |
| Sexual Abuse |
| Sexual Functioning |

What is your gender?

[ ] Male

[ ] Female

[ ] Transgender

[ ] Other: ___________________

What is your ethnicity? *Mark all that apply.*

[] Caucasian

[] Hispanic or Latino

[] African or African American

[] Native American

[] Asian or Pacific Islander

[] Other: ___________________________________

Do you have previous clinical experience taking a sexual history? If yes, please indicate the setting(s) in which this experience took place.

[ ] volunteering at a free clinic

[ ] previous standardized patient encounter through medical school

[ ] Other: _____________________________________

Were you born in the United States?

[] Yes

[] No

What is your year of training?

[ ] MS1

[ ] MS2
