## Appendix D for "“You have to add your own character”: Medical Student Perspectives on Observing Clinical Encounters"

**Appendix D. Interview Guide**

*I would like to ask you some questions now about your experiences today. This interview will be confidential and your participation is voluntary.*

1. How did you approach the standardized patient encounter? What was your plan when you entered the examination room? How did you develop that plan? What pitfalls did you try to avoid? What specific steps or behaviors did you want to make sure you executed?
2. What do you remember from the video? What particular aspects of the attending's performance did you pay attention to?
3. I'd like to talk with you about how you rated the encounter on this checklist. *(Review checklist)*
4. How well did the attending in the video do with taking a sexual history? What are some specific things she did well? What are some specific things she did poorly?
5. How did watching the video before the encounter influence your performance? Were there any specific things you saw the attending do that you wanted to replicate? Were there any specific things the attending did that you wanted to avoid?
6. How much do you relate to or identify with the attending in the video? What specific attributes influence your answer?

*Thank you for your participation!*

Positive Behaviors Modelled in the Video:

- Patient Confidentiality Assurance
- Good Listening Skills:
  - Seated next to the patient
  - Good eye contact
  - Periodically repeated patient's statements to assure understanding and reassure the patient that they are being heard
- Covered the "5 P's" recommended by the CDC for a sexual history to encompass risk factors for STIs and pregnancy
  - Partners
  - Practices
  - Protection from STIs
  - Past History of STIs
  - Prevention of Pregnancy

Negative Behaviors Modelled in the Video/Areas for improvement:

- Poor listening skills
  - Interruptions
- Use of gender exclusive terms
  - Always start by asking about partners, rather than assuming a specific gender
- Use of slang rather than anatomical/medical terms

    - For example: referring to his problem as “problems with erections” rather than “trouble getting it up”
- Ask more “normalizing” questions
    - preface STI question with “STIs are a common problem I see in my patients…” and re-phrase drug question to: “which drugs do you use?” In order to encourage disclosure of things that the patient might be unsure about telling his physician
- No inquiries about abuse/sexual abuse
    - “Has anyone ever forced you into sexual activities when you did not want them?” Or “Have you ever felt unsafe in your relationship?”
- Failure to acknowledge patient’s emotions
    - embarrassment at discussing ED, stress over marital strain from ED
